# Estimating the Effects of Vaping Frequency on Smoking Cessation Using PATH Study Data: A Causal Forest Approach

**DOI:** 10.64898/2026.09.02.26362051

**Authors:** Thuy T. T. Le, Erik Sverdrup, Ruoyan Sun, Irina Bondarenko, Mohammed Jaffri, Evelyn Jimenez-Mendoza, Jihyoun Jeon, Adam Leventhal, Andrew Hyland, Kenneth E. Warner, David Mendez

## Abstract

**Introduction:** E-cigarettes are the second most commonly used nicotine product in the United States after cigarettes. Their role in smoking cessation remains debated. Further research is needed to clarify their potential impact on quitting combustible cigarette use.

**Methods:** We analyzed data from adults aged 18 and older who participated in Wave 4 (12/2016-01/2018), Wave 5 (12/2018-11/2019), and Wave 7 (01/2022-04/2023) of the Population Assessment of Tobacco and Health Study. We employed a causal forest to estimate the associations between Wave 5 e-cigarette use and past 12-month cigarette smoking abstinence at Wave 7. Eligible participants had smoked at least 100 cigarettes in their lifetime at Wave 4, currently smoked cigarettes some days or every day at both Waves 4 and 5, completed follow-up at Wave 7, and had a substantial estimated probability of either vaping or not vaping at Wave 5. Analyses adjusted for 26 Wave 4 covariates, including demographic characteristics, tobacco use history, and other relevant factors.

**Results:** Compared with no vaping, vaping on 1-5 days was associated with a 2.4 percentage-point lower probability of cessation (95% CI: -7.7 to 2.9), whereas vaping on 6-29 days and daily vaping were associated with increases of 7.5 percentage points (95% CI: -0.4 to 15.5) and 12.5 percentage points (95% CI: 6.2 to 18.7), respectively.

**Conclusions:** The association between vaping and smoking cessation varied by vaping frequency. Compared with non-vaping, infrequent vaping was not significantly associated with subsequent cigarette abstinence, whereas more frequent vaping was associated with a greater likelihood of abstinence. Daily vaping showed the strongest positive association.

**Implications:** This study contributes to the ongoing debate about the role of e-cigarettes in smoking cessation. The findings suggest that vaping frequency may be an important factor in understanding whether e-cigarette use promotes smoking cessation, with more frequent vaping showing stronger associations with subsequent abstinence. Future research should further examine the conditions under which e-cigarette use may facilitate cessation and assess potential heterogeneity across population subgroups.

## Introduction

Cigarette smoking remains the leading cause of preventable disease, disability, and premature death in the United States (US), with approximately 480,000 smoking-attributable deaths per year [1]. Quitting smoking is well-known to be the most effective way to reduce mortality and morbidity among individuals who smoke cigarettes. However, achieving sustained smoking cessation remains a public health challenge. The rate of successful long-term quitting is low relative to the rate of making a quit attempt (about 9% vs. 53% in 2022, according to the National Health Interview Survey (NHIS) [2]).

E-cigarettes have been the second most commonly used nicotine product in the US, following cigarettes, with prevalence estimates of 7.0% and 9.9%, respectively, based on the 2024 NHIS [3]. Despite their widespread use, the role of e-cigarettes in smoking cessation remains a subject of ongoing debate. While not yet an FDA-approved cessation aid, using e-cigarettes as a substitute for cigarettes is the most popular way to quit smoking among US adults [4, 5]. A 2025 Cochrane review [6] synthesized evidence from randomized trials comparing e-cigarettes with control conditions, as well as uncontrolled intervention studies in which all participants received e-cigarettes. The review found that nicotine-containing e-cigarettes improved smoking cessation compared with nicotine replacement therapy, non-nicotine e-cigarettes, and behavioral support or no support, yielding approximately 3-4 additional quitters per 100 individuals who smoke cigarettes across comparisons.

Randomized controlled trials are considered the gold standard for evaluating the causal effect of e-cigarettes on smoking cessation. However, this approach is costly and may not fully capture the diversity and characteristics of the broader population of people who smoke. More importantly, randomized controlled trials may not reflect what happens under real-world conditions – that is, in the absence of the instruction and supervision that are often part of such trials. As such, further investigating this relationship using available observational data, such as the Population Assessment of Tobacco and Health (PATH) Study, could substantially complement existing evidence and deepen our understanding of this critical public health question. Several studies have applied causal inference methods to evaluate the effect of e-cigarette use on smoking cessation using the PATH data, with mixed findings reported, including both positive and negative associations; see [7] and references therein. Two studies conducted by Chen et al. [8, 9] used propensity score matching and difference-in-differences methods to estimate the association of e-cigarette use with subsequent smoking cessation among people who smoke with a recent quit attempt. Both studies applied the same analytic approach to different cohorts of survey participants who smoke in the PATH Study and found no evidence that vaping appeared to aid smoking cessation. Another study by Quach et al. [10], which also used the PATH Study and the same approach as Chen et al. [8, 9] to estimate the impact of daily and non-daily vaping at Wave 4 on smoking cessation at Wave 6 among Wave 4 participants who smoke, found that neither daily nor non-daily vaping was positively associated with smoking cessation. Another study by Harlow et al. [11] used marginal structural models, a causal inference method, to analyze PATH data across multiple waves. The study found that consistent daily vaping was positively associated with cigarette smoking abstinence, whereas consistent non-daily vaping was negatively associated. A 2026 review led by Xu [7] synthesized the findings from prior PATH-based studies on the effectiveness of nicotine vaping products for smoking cessation and found that 63% reported a positive association.

While causal machine learning methods have been developed and applied to estimate causal effects in many observational settings [12], their application to studying the impact of e-cigarette use on smoking cessation remains largely unexplored. To help address this gap and further strengthen the evidence on the relationship between vaping and smoking cessation, this study applied a causal forest [13] to estimate the association of e-cigarette use at Wave 5 on smoking cessation at Wave 7, using PATH data. Causal forests provide a flexible, data-driven approach to causal inference. They can adjust for a large set of measured confounders while capturing nonlinear relationships and complex interactions without requiring these features to be prespecified. The forest-based average exposure effect was estimated using a doubly robust procedure [14], allowing valid inference under appropriate regularity conditions when flexible machine learning methods, such as random forests, are used to model the outcome and exposure [15]. We focused on participants who had smoked at least 100 cigarettes in their lifetime by Wave 4, reported smoking some days or every day at both Waves 4 and 5, were followed up at Wave 7, and had an estimated propensity score indicating a substantial probability of either vaping or not vaping at Wave 5. We adjusted for a comprehensive set of 26 covariates measured at Wave 4. The exposure variable, past 30-day e-cigarette use, was measured at Wave 5, and the outcome variable, 12-month smoking abstinence, was measured at Wave 7. The findings of this study will contribute to a better understanding of whether vaping supports smoking cessation.

## Materials and Methods

### Data

The PATH study is an ongoing, nationally representative longitudinal survey that collects individual-level information to study tobacco and nicotine use and its health effects among individuals aged 12 and older in the US. Data have been collected annually or biannually, with the first wave in 2013. To date, data from eight main waves have been released publicly.

#### Analytic population

We analyzed a cohort of 3772 participants aged 18 and older who had smoked at least 100 cigarettes in their lifetime by Wave 4, smoked some days or every day at both Waves 4 (12/2016-01/2018) and 5 (12/2018-11/2019), and were followed up at Wave 7 (01/2022-04/2023).

#### Exposure Variable

The exposure variable was the number of days of e-cigarette use in the past 30 days at Wave 5. We examined the association between vaping frequency and smoking cessation by specifying the exposure variable in three ways: a four-category variable, a three-category variable, and a continuous variable. Based on prior evidence suggesting that infrequent vaping, defined as use on 5 or fewer days, may reflect curiosity or experimentation rather than motivation to quit smoking [16], we first categorized past 30-day e-cigarette use into four groups: 0 days, 1-5 days, 6-29 days, and 30 days. This categorization allowed us to compare smoking cessation among participants who vaped on 1-5, 6-29, or 30 days with smoking cessation among those who reported no vaping in the past 30 days. Second, we categorized e-cigarette use as no vaping (0 days), non-daily vaping (1-29 days), or daily vaping (30 days) and compared the associations of non-daily and daily vaping with smoking cessation relative to no vaping. Finally, we modeled the number of vaping days in the past 30 days as a continuous exposure variable.

#### Outcome Variable

The outcome of interest was past 12-month smoking abstinence status at Wave 7. Participants were classified as having past 12-month smoking abstinence if they reported not smoking any cigarettes during the 12 months preceding Wave 7.

#### Covariates

To estimate the association of e-cigarette use at Wave 5 with past 12-month smoking abstinence at Wave 7, we adjusted for a comprehensive set of 26 covariates measured at Wave 4. These variables include factors that are likely to influence both vaping and smoking cessation behaviors. Specifically, we included the following Wave 4 variables: age, sex, race, ethnicity, education level, smoking frequency (every day or some days), time to first nicotine product after waking, tobacco dependence score, average number of cigarettes smoked per day, menthol cigarette use, number of quit attempts, intention to quit, use of other tobacco products, e-cigarette use status, alcohol or other drug use, self-perceived physical and mental health, chronic disease status, health insurance status, perceived relative harm of e-cigarettes, perceived harm of cigarettes, time spent in close contact with people who smoke, home smoking rules, and exposure to e-cigarette marketing. Detailed descriptions of these variables are provided in the Appendix.

After merging Wave 4 data containing the selected covariates, Wave 5 data containing the exposure variable, and Wave 7 data containing the outcome variable, we excluded individuals with missing values, resulting in a final complete-case analytic sample of 3,772.

### Statistical Analysis

We estimated the average effect of vaping frequency on smoking cessation by three categorical vaping-frequency groups with a no-vaping reference group. Specifically, we estimated differences in the probability of smoking cessation in which all eligible participants vaped on 1-5 days, 6-29 days, or 6-30 days in the past 30 days, compared with all eligible participants who did not vape.

We first fitted a causal forest [13], a generalized random forest method that extends Breiman’s random forests [17] and is designed to identify heterogeneity in treatment or exposure effects across individuals [13]. We then estimated average exposure effects using an augmented inverse probability weighted estimator, with nuisance components, including the outcome and exposure models, obtained from the fitted forest.

Under standard causal identification assumptions [18], including consistency, conditional exchangeability (or no unmeasured confounding), and overlap, causal forests can be used to estimate the causal effect of vaping on smoking cessation. Of these assumptions, the overlap is the only one that can be empirically assessed using the observed data. The overlap assumption requires that, within strata of measured covariates, individuals have a nonzero probability of being in each exposure group. In observational studies, this assumption is not guaranteed because exposure is not randomized, and some covariate profiles may be strongly associated with vaping status. To address potential violations of positivity, we focused on the overlap population, which included individuals in the entire analytic population who had an estimated propensity score indicating a substantial probability of either vaping or not vaping at Wave 5 [19]. The overlap population represents a reweighted version of the original study population. Rather than excluding individuals in regions of limited overlap, overlap weighting assigns greater weight to individuals with high probabilities of belonging to the vaping and non-vaping groups and less weight to those with a strong propensity toward either group [19]. Thus, the original sample is retained, but the resulting overlap weights define the target population. If overlap violations are minimal, estimates in the overlap population would be expected to be similar to estimates in the full analytic population of 3772 participants, as defined in the previous section.

Causal forests flexibly adjust for measured confounders while accommodating nonlinear relationships and interactions among covariates. The forest-based average exposure effect estimator was constructed using a doubly robust approach [14], which enables valid inference when the outcome and exposure models are estimated with flexible machine learning models, such as random forests [15]. To account for the complex design of the PATH Study, we incorporated the Wave 7 all-wave longitudinal weights through the *sample weights* argument in *causal forest*. Cluster identifiers were constructed using the PATH pseudo-strata and pseudo-primary sampling unit variables and incorporated through the *clusters* argument. Variance estimates were calculated using Wave 7 replicate weights with balanced repeated replication and Fay’s adjustment, with ρ = 0.3 [20]. Analyses were conducted using the *causal forest* function in the *grf* package [21].

## Results

### Characteristics of the analytic sample

The analytic sample included 3,772 individuals from Waves 4 and 5 who reported having smoked at least 100 cigarettes in their lifetime, currently smoked cigarettes some days or every day at both waves, and completed follow-up at Wave 7. The sample included 1372 (34.2%) participants aged 18-34 years, 1501 (41.0%) aged 35-54 years, and 899 (24.8%) aged 55 years or older. Among these participants, 1673 (52.6%) were male, and 2099 (47.4%) were female. The racial/ethnic composition of the sample was 450 (11.3%) Hispanic participants, 2391 (68.8%) non-Hispanic White participants, 677 (14.6%) non-Hispanic Black participants, and 254 (5.4%) participants of non-Hispanic other races/ethnicities. A complete description of the analytic sample characteristics is provided in Table 1.

**Table 1:**
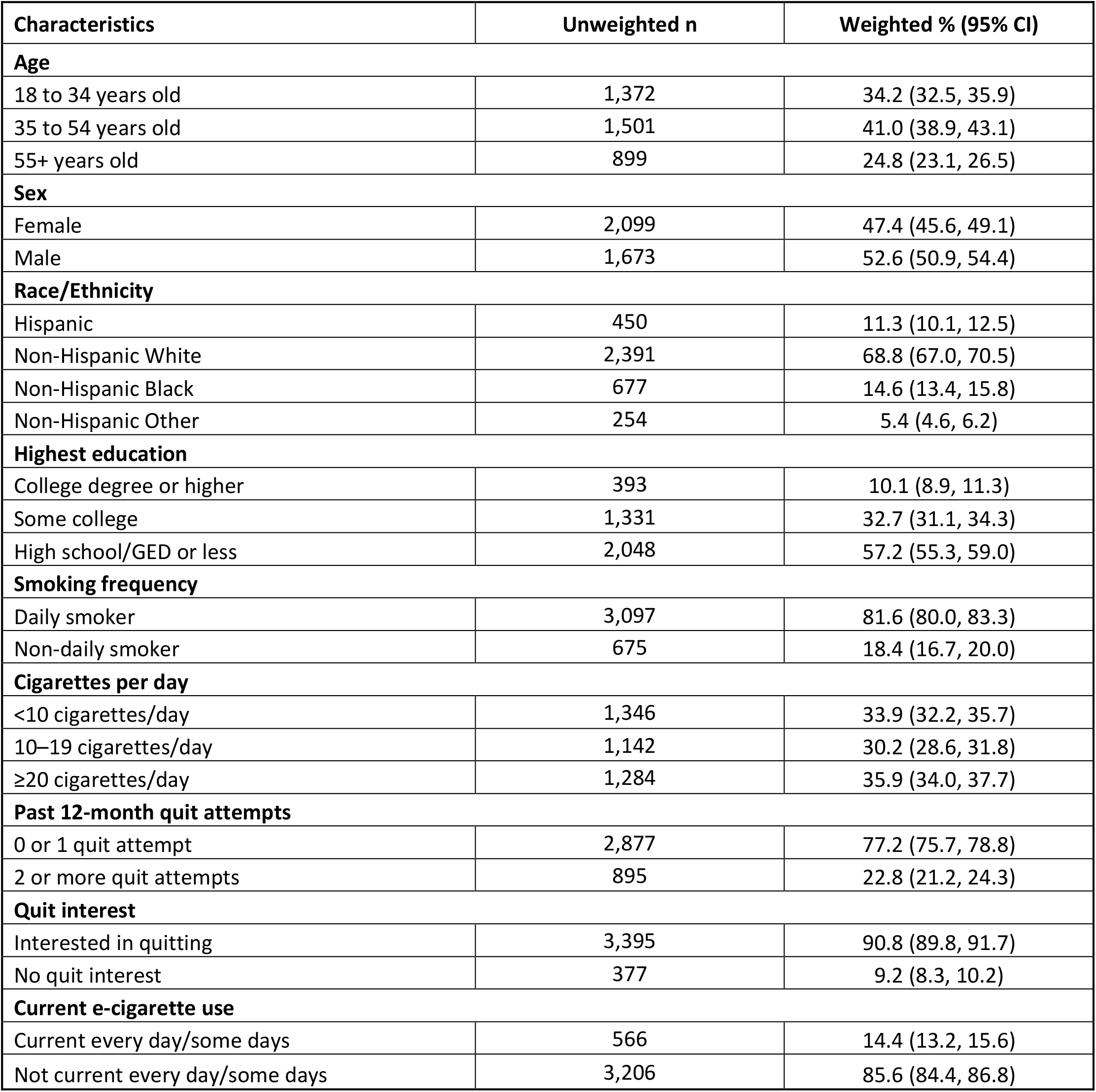
Baseline characteristics of the analytic sample (n = 3,772).

At Wave 5, 609 participants reported using e-cigarettes in the past 30 days. At Wave 7, 392 participants reported no cigarette smoking in the past 12 months. Appendix Table A1 presents the distribution of e-cigarette use days at Wave 5 by Wave 7 smoking status, with each reported number of days treated as a distinct value.

### Comparing the effects of different frequencies of e-cigarette use in the past 30 days at Wave 5

Table 2 shows that, in the overlap population, vaping frequency at Wave 5 was associated with smoking cessation by Wave 7. Compared with participants who reported no vaping, those who reported vaping on 1-5 days of the past 30 days had an estimated 2.4 percentage-point lower cessation rate, although the 95% confidence interval included zero, indicating statistical uncertainty. Those who reported vaping on 6-29 days had an estimated 7.5 percentage-point higher cessation rate (95% CI: -0.4 to 15.5), but this confidence interval also included zero. Participants who reported daily vaping (30 days) had the highest estimated cessation rate, which was 12.5 percentage points higher than that among participants who reported no vaping (95% CI: 6.2 to 18.7). Results for the full analytic population are presented in Appendix Table A2.

**Table 2:**
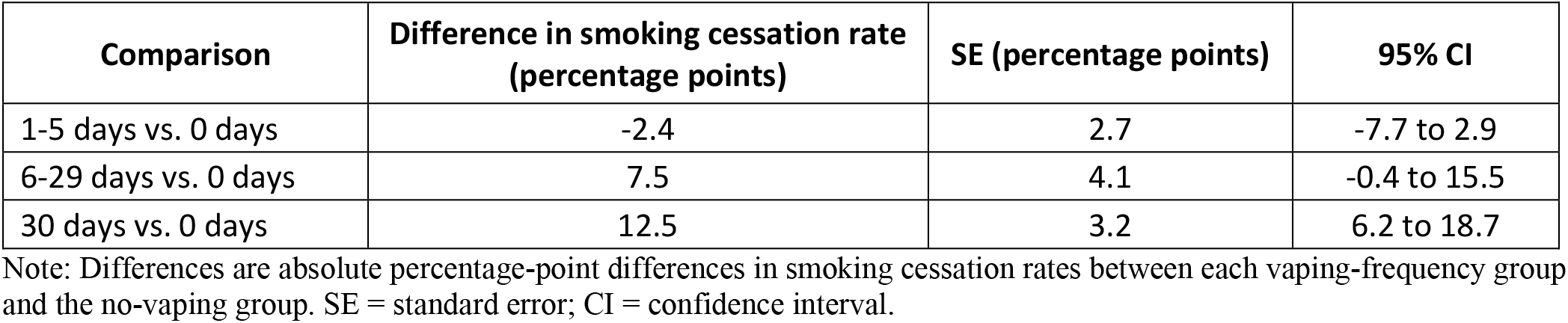
Estimated percentage-point differences in smoking cessation at Wave 7 by past 30-day vaping frequency at Wave 5 (1-5, 6-29, or 30 days vs. 0 days) in the overlap population.

| Comparison | Difference in smoking cessation rate (percentage points) | SE (percentage points) | 95% CI |
| --- | --- | --- | --- |
| 1-5 days vs. 0 days | -2.4 | 2.7 | -7.7 to 2.9 |
| 6-29 days vs. 0 days | 7.5 | 4.1 | -0.4 to 15.5 |
| 30 days vs. 0 days | 12.5 | 3.2 | 6.2 to 18.7 |

Note: Differences are absolute percentage-point differences in smoking cessation rates between each vaping-frequency group and the no-vaping group. SE = standard error; CI = confidence interval.

As seen in Table 3, compared with no vaping, non-daily vaping was associated with a non-significant 2.0 percentage point increase in the likelihood of smoking cessation (95% CI: -2.9 to 6.9 percentage points). When the number of vaping days in the past 30 days at Wave 5 was modeled as a continuous exposure, each additional vaping day was associated with a 0.44 percentage-point increase in the probability of smoking cessation (95% CI: 0.24 to 0.64 percentage points). These estimates are presented in Table 3 and Appendix Table A4. (See Appendix Tables A3-A4 for the results of these analyses conducted in the entire analytic population.)

**Table 3:** Estimated percentage-point differences in smoking cessation at Wave 7 by past 30-day vaping frequency at Wave 5 (1-29, or 30 days vs. 0 days) in the overlap population.

| Comparison | Difference in smoking cessation rate (percentage points) | SE (percentage points) | 95% CI |
| --- | --- | --- | --- |
| 1-29 days vs. 0 days | 2.0 | 2.5 | -2.9 to 6.9 |
| 30 days vs. 0 days | 12.5 | 3.2 | 6.2 to 18.7 |

## Discussion

This study used a causal forest to estimate the association between vaping frequency at Wave 5 and past 12-month smoking cessation at Wave 7 in the overlap population. This population included participants who completed Waves 4 and 5, had smoked at least 100 cigarettes in their lifetime, currently smoked cigarettes some days or every day at both waves, completed follow-up at Wave 7, and had a substantial probability of either vaping or not vaping at Wave 5. In this population, we compared the smoking cessation rate among participants who reported no vaping with the rates among those who reported vaping at various frequencies in the past 30 days at Wave 5. These analyses were adjusted for a comprehensive set of Wave 4 covariates – including age, race, education level, motivation to quit, prior quit attempts, and other covariates.

Compared with no vaping, infrequent vaping (1-5 days in the past 30 days) was negatively associated with smoking cessation, although the estimate was statistically uncertain. More frequent e-cigarette use at Wave 5 was generally associated with a higher likelihood of past-12-month smoking cessation at Wave 7. Specifically, daily vaping was associated with the largest estimated increase in smoking cessation among the vaping-frequency categories examined (Table 2). Furthermore, there was no evidence that, on average, nondaily vaping (1-29 days) was associated with a higher likelihood of smoking cessation than no vaping (Table 3). Analyses treating the number of vaping days in the past 30 days as a continuous exposure provided additional evidence of a positive association between vaping frequency at Wave 5 and past 12-month smoking cessation at Wave 7 (Appendix Table A4). Taken together, these findings suggest that the association between vaping and smoking cessation varies by vaping frequency, with more frequent vaping generally associated with a greater likelihood of cessation.

This study has several limitations. Interpretation of the findings as causal, rather than purely associative, depends on standard causal assumptions, including conditional exchangeability, overlap, and consistency. Although we adjusted for a comprehensive set of Wave 4 covariates selected based on subject-matter expertise, potential unmeasured confounding may still exist. Therefore, if these assumptions are not met, the estimates obtained from the causal forest should be interpreted as associations between vaping frequency and smoking cessation rather than as causal effects.

E-cigarette use is a heterogeneous exposure. Although vaping frequency was measured as the number of vaping days in the past 30 days at Wave 5, this measure does not capture device type, nicotine concentration, intensity of use, flavor, or reasons for vaping, or whether e-cigarettes were used specifically as a cessation aid. Therefore, people who smoke but report the same number of vaping days may have different experiences with vaping. Furthermore, because vaping exposure was measured at Wave 5, we could not fully capture changes in vaping behavior between Wave 5 and Wave 7, like other previous studies [8-10]. Some participants may have discontinued, initiated, or changed their frequency of e-cigarette use after Wave 5. As a result, these estimates reflect the associations of Wave 5 vaping frequencies rather than those of sustained vaping with Wave 7 smoking cessation over the follow-up period.

Smoking cessation and e-cigarette use were self-reported and may be subject to recall error or other biases [9, 22]. Loss to follow-up and missing data may introduce selection bias if participants retained through Wave 7 differed systematically from those lost to follow-up in ways related to both vaping and smoking cessation. Although survey weights may reduce this concern, attrition-related bias cannot be completely excluded.

This study introduced causal forests, a causal machine learning approach, for causal inference in tobacco control research. Although causal forests have been applied in other fields, their use in tobacco control remains limited [12]. This approach provides a flexible, data-driven method for handling a large set of covariates without requiring pre-specified relationships or interactions among them. The forest-based average exposure effect was estimated using augmented inverse probability weighting, yielding a doubly robust estimate that is less sensitive to model misspecification. However, because causal forests rely on stochastic procedures, the resulting estimates may vary slightly depending on random variation and hyperparameter tuning.

In summary, our findings indicate that the association of e-cigarette use with smoking cessation varies by vaping frequency. More frequent vaping was associated with a higher probability of achieving 12-month smoking abstinence, whereas infrequent vaping showed less evidence of supporting cessation. This observation suggests that sustained e-cigarette use may be more relevant for smoking cessation than intermittent or experimental use. A possible explanation is that frequent e-cigarette use provides people who smoke with a more robust alternative to cigarettes, which may help reduce withdrawal symptoms and lower dependence on combustible cigarettes, thus facilitating the transition away from smoking. In contrast, occasional vaping may not provide sufficient nicotine substitution or behavioral reinforcement to support sustained abstinence. These findings underscore the importance of e-cigarette use frequency in studies of the effect of vaping on smoking cessation. Our study’s findings add more evidence to the existing literature regarding the role of e-cigarettes in aiding smoking cessation [5-7].

## Data Availability

All data produced are available online at https://www.icpsr.umich.edu/web/NAHDAP/studies/36498/versions/V25

## Funding

T.T.T.L, I.B., M.J., E.J-M, J.J., K.E.W., and D.M. were supported by the National Cancer Institute of the National Institutes of Health and the Food and Drug Administration Center for Tobacco Products (Award Number 2U54CA229974). The content is solely the responsibility of the authors and does not necessarily represent the official views of the NIH or the Food and Drug Administration.

## Declaration of Competing Interests

None

## Notes

### Competing Interest Statement

The authors have declared no competing interest.

### Author Declarations

The study used ONLY publicly available data that were originally located at: https://www.icpsr.umich.edu/web/NAHDAP/studies/36498/versions/V25

