## Appendix for "Estimating the Effects of Vaping Frequency on Smoking Cessation Using PATH Study Data: A Causal Forest Approach"

### 1) <u>Description and derivation of 26 Wave 4 baseline covariates, Wave 5 exposure variable, and Wave 7 outcome variable.</u>

**Age**: Age was measured using the PATH Wave 4-derived age-category variable R04R_A_AGECAT6.

**Sex**: Sex was measured using the PATH Wave 4-derived sex variable R04R_A_SEX.

**Hispanic ethnicity**: Hispanic ethnicity was measured using the PATH Wave 4-derived Hispanic ethnicity variable R04R_A_HISP.

**Race**: Race was measured using the PATH Wave 4-derived three-category race variable R04R_A_RACECAT3.

**Education**: Education level was measured using the PATH Wave 4-derived education variable R04R_A_AM0018_V2.

**Cigarette smoking status**: Cigarette smoking frequency at Wave 4 was measured as smoking every day versus some days using R04_AC1003.

**Time to first nicotine product after waking**: Time to first nicotine product after waking was calculated in minutes using product-specific timing variables for cigarettes, e-cigarettes, cigars, pipes, snus, smokeless tobacco, and multiple-product use (R04_AC1024_NN/UN, R04_AV1024_NN/UN, R04_AG1024TC_NN/UN, R04_AG1024CG_NN/UN, R04_AG1024FC_NN/UN,

R04_AP1024_NN/UN, R04_AU1024_NN/UN, R04_AS1024_NN/UN, R04_AY0010_NN/UN). Hour-based responses were converted to minutes, and the earliest non-missing time across products was used.

**Menthol cigarette use**: Menthol cigarette use was coded as yes if participants reported menthol or mint cigarette use on any relevant Wave 4 item (R04_AC1130MC, R04_AC1130RY, R04_AC1050, R04_AC9150); otherwise, participants were coded as no menthol use.

**Tobacco dependence score**: Tobacco dependence was the PATH Wave 4 adult dependence score for current and dual users of non-electronic products and was derived using a graded response model on the combination of the Wave 4 variables R04_AN0055, R04_AN0025, R04_AN0030, R04_AN0035, R04_AN0045, R04_AN0060, R04_AN0065, R04_AN0050, R04_AN0070, R04_AN0075, R04_AN0080, R04_AN0085, R04_AN0090, R04_AN0095, R04_AN0100, R04_AN0813. This set of items is based on the publication by Strong et al. [1].

**Average number of cigarettes smoked per day**: The average number of cigarettes smoked per day was derived by combining reports from everyday smokers and someday smokers (R04R_A_PERDAY_EDY_CIGS, R04R_A_PERDAY_P30D_CIGS). The first non-missing value was used.

**Quit attempts**: The number of quit attempts was measured using R04_AN0115. Participants who reported no quit attempt in the past 12 months (R04_AN0105 = No) were assigned a value of zero.

**Intention to quit**: Intention to quit smoking was measured using the Wave 4 item R04_AN0230.

**Other combustible tobacco use:** The frequency of other combustible tobacco use included respondents who reported current every day or some days use of traditional cigars, cigarillos, filtered cigars, pipes, or hookah. To do this, we combined the PATH-derived variables R04R_A_CUR_EDSD_GTRAD, R04R_A_CUR_EDSD_GRILLO, R04R_A_CUR_EDSD_GFILTR, R04R_A_CUR_EDSD_PIPE, and R04R_A_CUR_EDSD_HOOK. Participants who reported current every day or some days use of any of these products were classified as other combustible tobacco users, and those who reported no use of any of these products were classified as non-users.

**Smokeless tobacco use**: The frequency of smokeless tobacco use included respondents who reported current every day or some days use of snus, or smokeless tobacco. Based on the PATH-derived variables R04R_A_CUR_EDSD_SNUS, and R04R_A_CUR_EDSD_SMKLS, participants who reported current every day or some days use of any of these products were classified as smokeless tobacco users, and those who reported no use of any of these products were classified as non-users.

**Current e-cigarette use**: Current every day or some day e-cigarette use was measured using the PATH Wave 4 derived variable R04R_A_CUR_EDSD_EPRODS.

**Alcohol or other drug use**: This variable was derived from the Wave 4 item R04_AX0170, which assessed the last time the respondent used alcohol or other drugs weekly or more often. Respondents who reported weekly or more frequent use in the "past month" or "2 to 12 months ago" were classified as having used alcohol or other drugs weekly or more in the past 12 months. All other respondents were classified as having no weekly or more frequent alcohol or other drug use in the past 12 months.

**Self-rated physical health**: Self-perception of physical health was measured using the Wave 4 variable R04_AX0090.

**Chronic disease history**: This variable was derived as a cumulative indicator of whether a respondent reported any chronic disease across Waves 1-4. For each wave, respondents were classified as having a chronic disease if they reported any marked heart condition, any marked respiratory condition, or a "Yes" response for cancer or diabetes. Survey variables used included heart condition items AX0111, respiratory condition items AX0119, cancer AX0144, and diabetes AX0281 from Waves 1-4, including Wave 2–4 new baseline (NB) and 12-month (12M) versions where applicable.

**Externalizing mental health symptoms**: Externalizing symptoms in the past 12 months were calculated by summing seven Wave 4 symptom items (R04_AX0165, R04_AX0166, R04_AX0167, R04_AX0168, R04_AX0169, R04_AX0250, R04_AX0251). Participants were categorized as low (1), moderate (2-3), or high (>=4) externalizing symptoms based on the number of symptoms reported.

**Internalizing mental health symptoms**: Internalizing symptoms in the past 12 months were calculated by summing four Wave 4 symptom items (R04_AX0161, R04_AX0162, R04_AX0163, R04_AX0164). Participants were categorized as low (1), moderate (2-3), or high (>=4) internalizing symptoms based on the number of symptoms reported.

**Health insurance**: Health insurance status was measured using the PATH Wave 4-derived indicator of health insurance coverage R04R_A_AM0026_V2.

**Perceived harm of e-cigarettes**: Perceived harmfulness of e-cigarettes or other electronic nicotine products compared with cigarettes was measured using R04_AE1099.

**Perceived harm of cigarettes**: Perceived harmfulness of cigarettes to health was measured using R04_AC9050.

**Secondhand smoke exposure**: Exposure to others' smoking was measured as the number of hours in the past 7 days that participants were in close contact with others while they were smoking (R04_AX0068).

**Home smoking rules**: Rules about smoking combustible tobacco products inside the home were measured using R04_AR1045.

**ENDS marketing exposure**: Exposure to e-cigarette or electronic nicotine product marketing was coded as exposed if participants reported any ENDS-related advertisement exposure across media sources (R04_AX0203_02-R04_AX0203_10) or exposure to ENDS-related promotions, coupons, or free samples (R04_AX0678, R04_AX0708_02). Otherwise, participants were coded as not exposed.

**Number of days using e-cigarettes in the past 30 days at Wave 5**: The number of days a respondent used ENDS in the past 30 days was coded as zero if the participant reported that they did not use ENDS in the past 30 days (R05_AV1004), in the past 12 months (R05_AV1002_12M), or that they have not used ENDS even once or twice (R05_AV1002), or that they now use ENDS "not at all" (R05_AV1003). The number of days was coded as the reported value when the respondent provided a number of days they used ENDS in the past 30 days (R05_AV1022). The number of days was coded as 30 if the respondent reported using ENDS "every day" (R05_AV1003).

**Past 12-month smoking status at Wave 7**: Past 12-month smoking status at Wave 7 was measured using the Wave 7 derived-variable R07R_A_P12M_CIGS.

### 2) Estimated Propensity Scores

The estimated propensity scores ranged from 0.014 to 0.5, suggesting that most participants had a nonzero estimated probability of being in either the vaping or no-vaping group, as shown in Figure A1. After weighting, measured covariates were well balanced across groups, as shown in Figures A2.A-C. These results indicate that the overlap assumption was not severely violated. Nevertheless, a small subset of participants had estimated propensity scores close to zero, suggesting limited overlap for certain covariate profiles. This limited overlap may explain the differences in the estimates observed in the full analytic population presented in Appendix Tables A2-A4, compared with those observed in the overlap population.

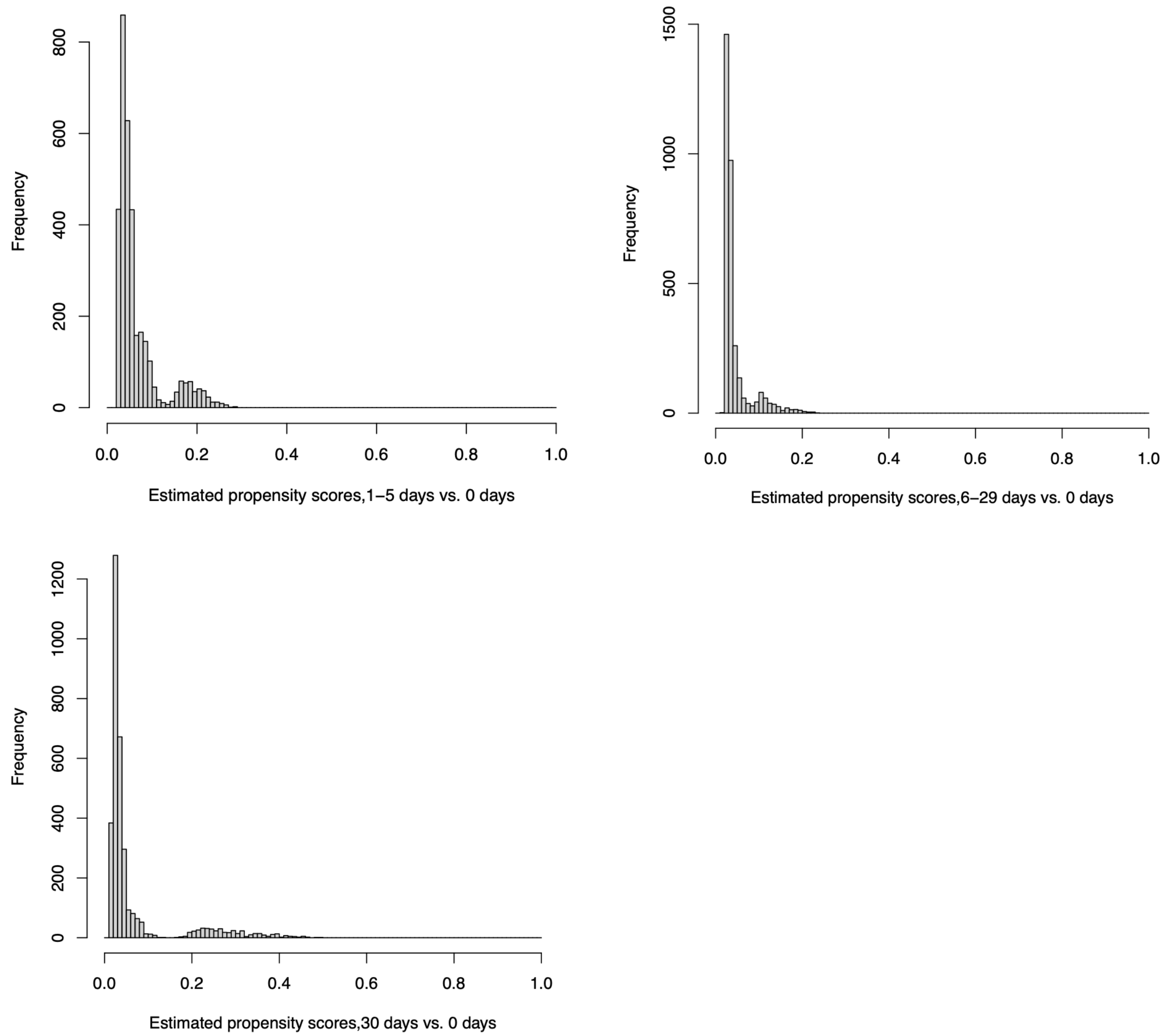


**Figure A1**: Histograms of the estimated propensity scores.

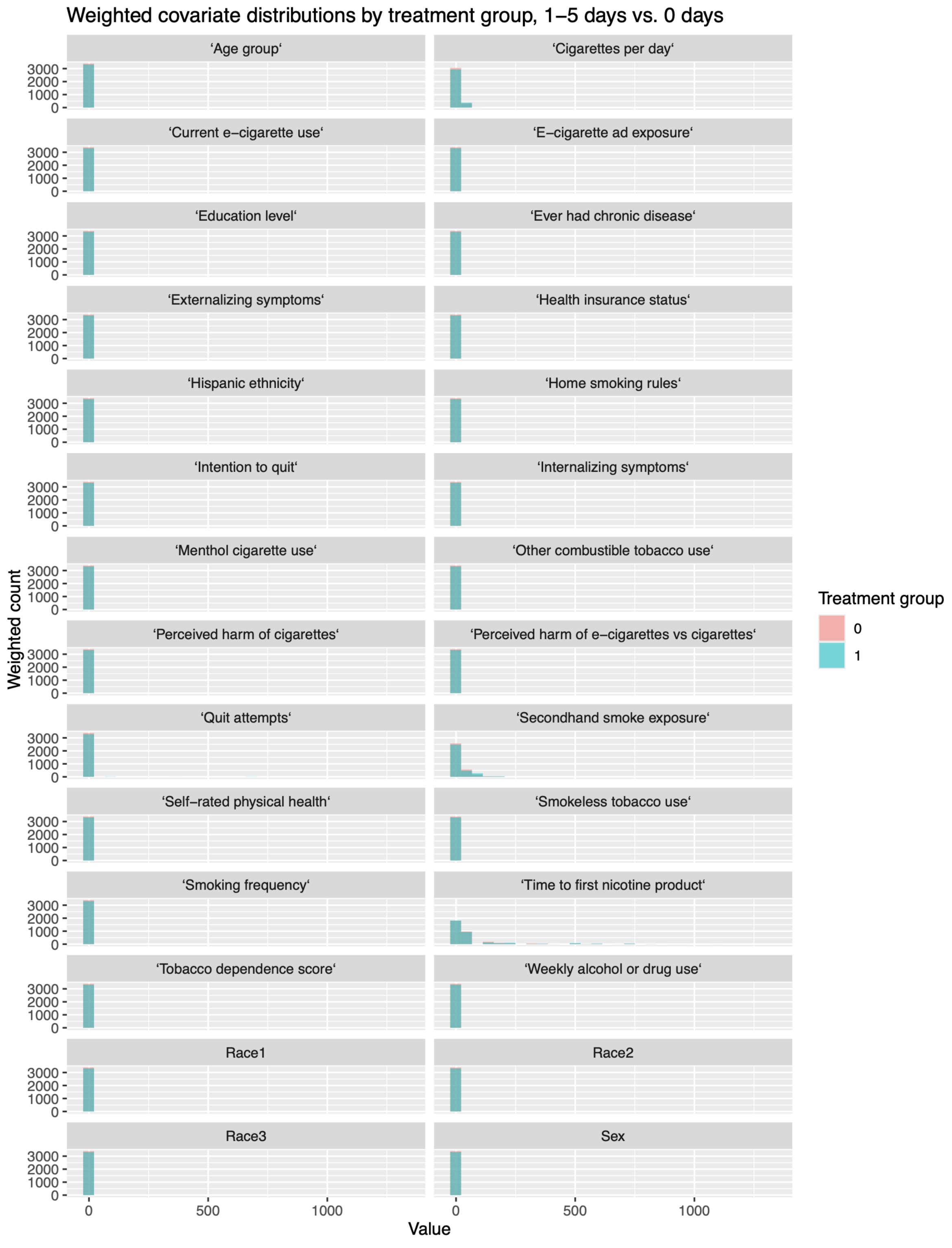


**Figure A2.A**: Distribution of weighted covariate values by treatment group (1-5 days vs. 0 days).

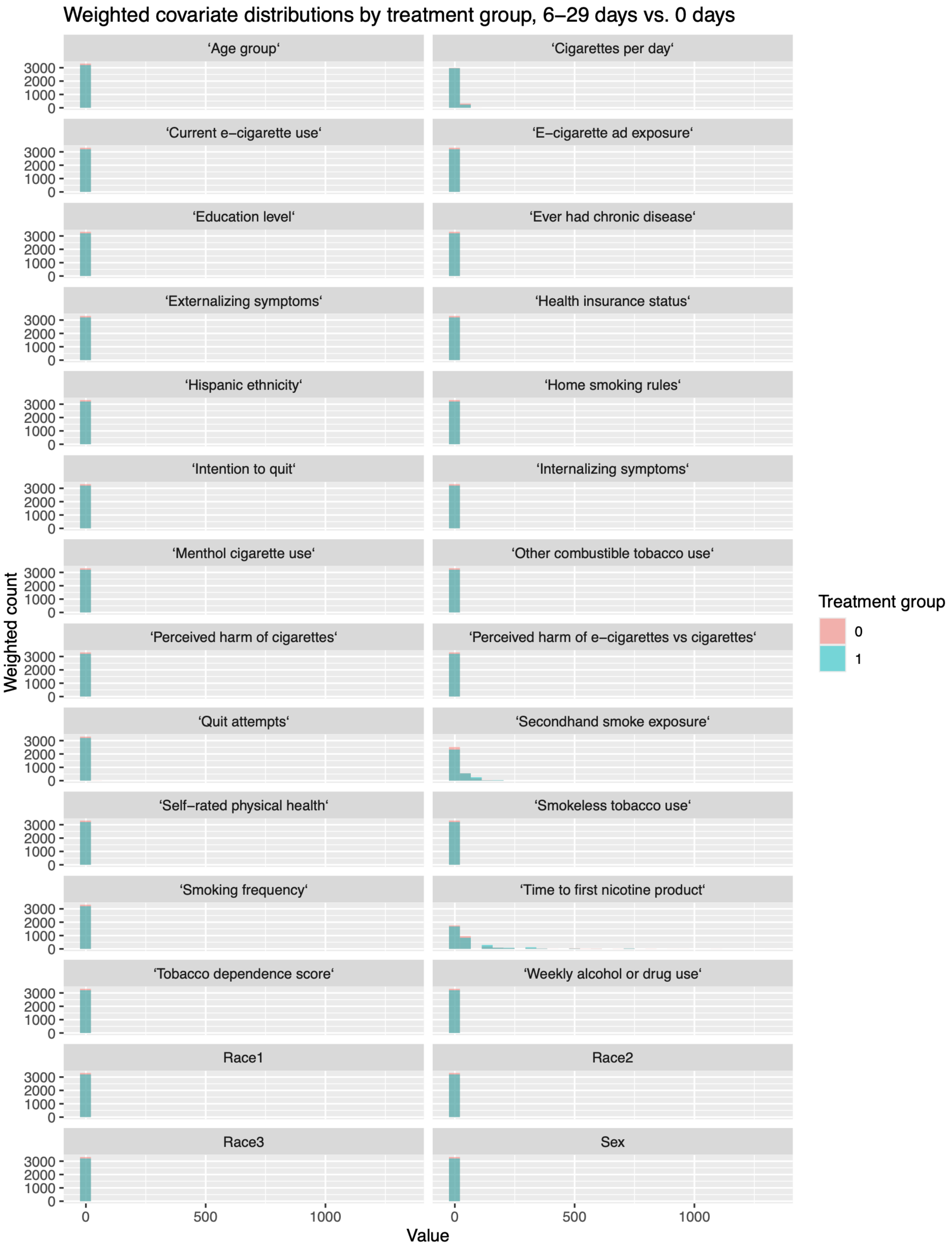


**Figure A2.B**: Distribution of weighted covariate values by treatment group (6-29 days vs. 0 days).

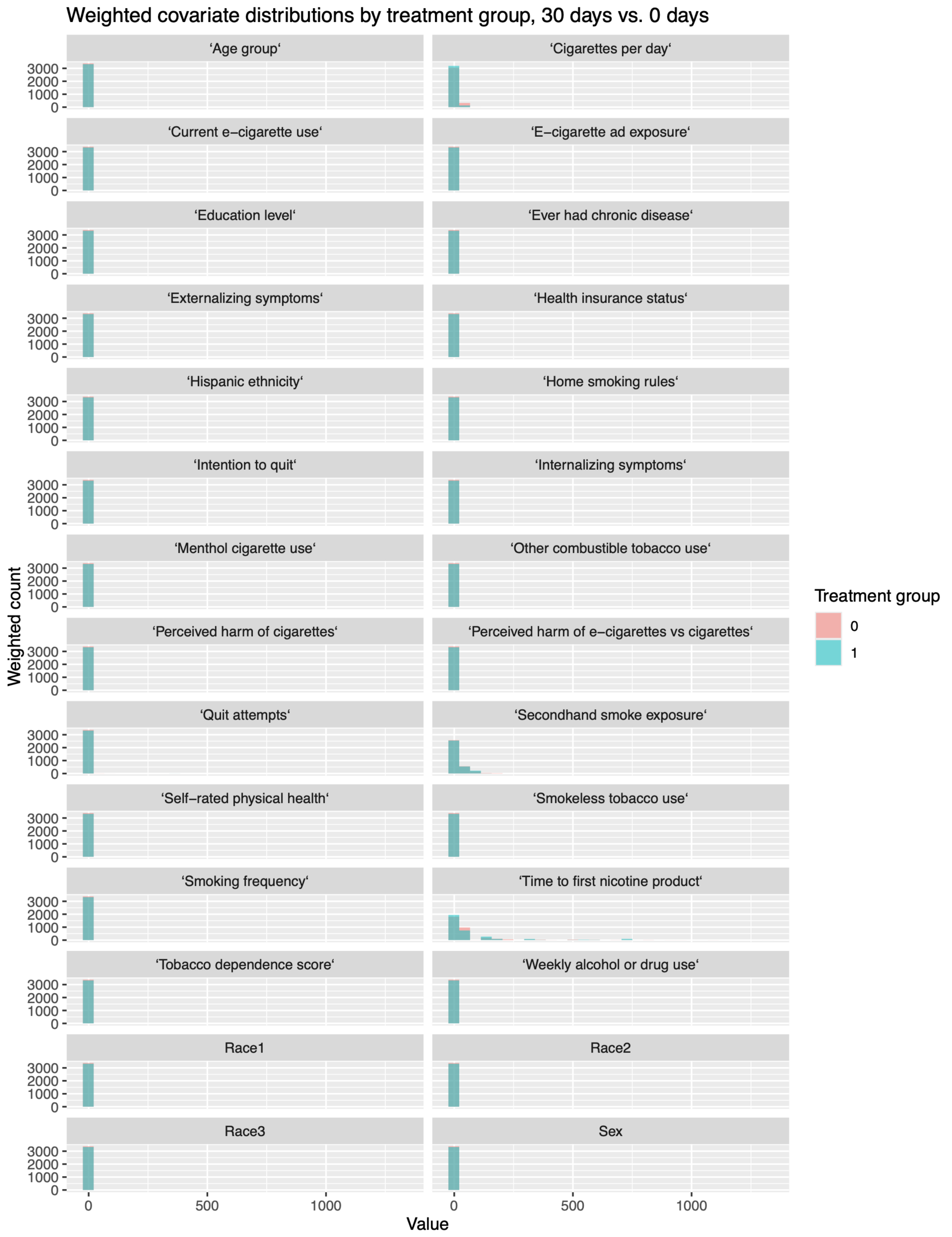


**Figure A2.C**: Distribution of weighted covariate values by treatment group (30 days vs. 0 days).

### 3. Additional analysis

| Number of e-cigarette use days | No past 12-month smoking abstinence | | Past 12-month smoking abstinence | |
|---|---|---|---|---|
| | Unweighted n | Unweighted % | Unweighted n | Unweighted % |
| 0 | 2861 | 84.6% | 302 | 77.0% |
| 1 | 51 | 1.5% | 3 | 0.8% |
| 2 | 67 | 2.0% | 5 | 1.3% |
| 3 | 40 | 1.2% | 6 | 1.5% |
| 4 | 10 | 0.3% | 0 | 0.0% |
| 5 | 53 | 1.6% | 1 | 0.3% |
| 6 | 9 | 0.3% | 2 | 0.5% |
| 7 | 2 | 0.1% | 0 | 0.0% |
| 8 | 10 | 0.3% | 1 | 0.3% |
| 9 | 5 | 0.1% | 0 | 0.0% |
| 10 | 33 | 1.0% | 6 | 1.5% |
| 11 | 3 | 0.1% | 0 | 0.0% |
| 12 | 5 | 0.1% | 1 | 0.3% |
| 13 | 1 | 0.0% | 1 | 0.3% |
| 14 | 2 | 0.1% | 0 | 0.0% |
| 15 | 24 | 0.7% | 5 | 1.3% |
| 16 | 1 | 0.0% | 0 | 0.0% |
| 17 | 0 | 0.0% | 1 | 0.3% |
| 18 | 2 | 0.1% | 0 | 0.0% |
| 20 | 20 | 0.6% | 3 | 0.8% |
| 25 | 9 | 0.3% | 2 | 0.5% |
| 26 | 1 | 0.0% | 0 | 0.0% |
| 28 | 5 | 0.1% | 0 | 0.0% |
| 29 | 1 | 0.0% | 0 | 0.0% |
| 30 | 165 | 4.9% | 53 | 13.5% |

**Table A1:** Distribution of past 30-day e-cigarette use days at Wave 5 by past 12-month smoking abstinence at Wave 7 in the entire analytic population.

| Comparison | Difference in smoking cessation rate (percentage points) | SE (percentage points) | 95% CI |
|---|---|---|---|
| 1-5 days vs. 0 days | -1.2 | 3.4 | -7.8 to 5.3 |
| 6-29 days vs. 0 days | 12.4 | 6.9 | -1.0 to 25.8 |
| 30 days vs. 0 days | 10.2 | 5.4 | -0.5 to 20.9 |

**Table A2**: Estimated percentage-point differences in smoking cessation at Wave 7 by past 30-day vaping frequency at Wave 5 (1-5, 6-29, or 30 days vs. 0 days) in the full analytic population.

| Comparison | Difference in smoking cessation rate (percentage points) | SE (percentage points) | 95% CI |
|---|---|---|---|
| 1-29 days vs. 0 days | 5.3 | 3.8 | -2.2 to 12.8 |
| 30 days vs. 0 days | 10.2 | 5.4 | -0.5 to 20.9 |

**Table A3**: Estimated percentage-point differences in smoking cessation at Wave 7 by past 30-day vaping frequency at Wave 5 (1-29, or 30 days vs. 0 days) in the full analytic population.

| Population | Estimated increase in smoking cessation rate per additional day of vaping (percentage points) | SE (percentage points) | 95% CI |
|---|---|---|---|
| Entire analytic Population | 0.36 | 0.22 | -0.07 to 0.78 |
| Overlap population | 0.44 | 0.10 | 0.24 to 0.64 |

**Table A4**: Estimated change in smoking cessation rate associated with one additional day of vaping. Values are reported in percentage points with standard errors (SE) and 95% confidence intervals (CIs).
